# Gastrointestinal and Fluid Retention Symptoms Associated with Creatine Monohydrate With and Without Loading Dose Over 28 Days of Supplementation

**DOI:** 10.1101/2025.10.07.25337280

**Authors:** Jon C Wagner, Mark Faulkner, William Faulkner, Fredrick Mullins

## Abstract

**Background:** Creatine monohydrate (CM) is widely used for improving athletic performance and supporting general health. Despite its well-established safety profile, gastrointestinal (GI) distress and varying issues related to fluid retention remains a commonly reported side effect, particularly at higher doses. As CM is increasingly utilized in non-athletic populations—including females who are underrepresented in performance trials and patients with a variety of medical conditions that may require larger doses—understanding the tolerability of different dosing regimens is essential.

**Objective:** This study aimed to investigate the incidence and severity of GI symptoms and fluid retention associated with daily CM supplementation, with and without a loading phase, in healthy male and female adults.

**Methods:** In a 28-day single-center, single-blind, randomized clinical trial conducted by Princeton Consumer Research Corp., 24 healthy participants were assigned to either Group A (5 g/day CM) or Group B (20 g/day loading dose for 14 days followed by 5 g/day maintenance). Blood and urine samples were collected at baseline and day 28 to assess safety. GI symptoms were recorded via self-report using a structured severity rating scale.

**Results:** All participants completed the study without any serious adverse events. One non-serious adverse event (menstrual cramps) was reported and deemed unrelated to CM use. Undesired GI symptoms were reported by 79.2% of all participants and by 81.0% of females, with bloating, water retention, puffiness, and stomach discomfort being the most frequently cited. A higher proportion of participants in the loading dose group (Group B) reported GI symptoms, which were also rated as more severe compared to the standard dose group (Group A), though differences did not reach statistical significance. Laboratory analyses revealed no clinically significant changes, indicating both regimens were safe over 28 days.

**Conclusion:** CM supplementation at both standard and loading doses is generally safe but frequently associated with mild to severe GI symptoms. While not statistically significant, a trend toward greater frequency and severity of symptoms in the loading group suggests a potential dose-dependent effect. These findings warrant further research on higher CM doses, tolerability in diverse populations, and whether more soluble creatine formulations may reduce GI side effects and improve adherence.

## Introduction

Creatine supplementation has been a common method to improve exercise and athletic performance since the early 1990s. (1,2) Since then creatine has become popular with the general population as an adjunct for fitness performance and health. (3,4,5) Although generally regarded as safe and effective, anecdotal reports persist linking creatine, particularly given as the monohydrate salt (CM), with gastrointestinal (GI) distress and water retention. One study compared GI adverse events at two dosing levels to placebo in professional male soccer players. They showed a significant difference, specifically for diarrhea, in the higher dose of CM compared to the lower CM dose suggesting a dose dependent effect. (6) These results have two implications. First, as more pre and perimenopausal females consider creatine supplementation the issue of GI distress and fluid retention may be viewed as a negative. Secondly, creatine is being studied for health purposes at doses substantially higher than what is currently used by athletes. (7,8,9) If GI symptoms and bloating are dose dependent phenomenon then the larger doses of CM could negatively impact adherence.The objective of this study was to investigate the effects on well-being of creatine monohydrate supplementation given with and without a loading dose on the incidence of GI symptoms and fluid retention in male and female subjects.

## Methods

### Subjects

This trial was a single center, single blind, randomized clinical study design over twenty-eight days. The study was conducted at Princeton Consumer Research Corp. located in Raritan, New Jersey between May 30, 2024 and July 3, 2024. The study (IRB2024-SP029) was approved by the UNIVO investigational review board on May 2, 2024 and registered on clinicaltrials.gov (NCT07176325).

#### Inclusion criteria

○ Subjects must be healthy males or females, aged 18 to 60 inclusive at the time of screening.
○ Subjects must be able to understand the study, agree to the requirements and restrictions, and be willing to give voluntary consent to participate in the study.
○ Subjects are willing to have up to two (2) blood collections, one at Visit 2 and Visit 3.
○ Subjects are willing to provide a urine sample that is collected over 24 hours. This entails collecting all urine in a 24-hour period in a container.
○ Subject is willing to fast for 12 hours before the Baseline visit (second visit). Water is permitted.
○ Subject is willing to have waist measurements taken with calipers and willing to wear clothing that allows easy access to their waist.
○ Female subjects of childbearing potential must agree to one of the following methods of birth control through the duration of the study. Acceptable methods include: abstinence, same-sex partner, double barrier (condom, diaphragm, or cervical cap with spermicidal foam, gel, or cream); intra-uterine device (IUD) with or without hormones in place or hormonal contraception (oral, injectable, implantable, transdermal, or vaginal) used consecutively for at least 3 month prior to screening visit; vasectomized partner or bilateral insertion of Essure® implants (or analogous) for at least 6 months prior to the screening visit; to be considered a female of non-childbearing potential, subject must have had a bilateral tubal ligation, hysterectomy or bilateral oophorectomy; or postmenopausal status with amenorrhea (no menstruation) for at least 1 year prior to the screening visit.
○ Subject is willing to comply with the study restrictions.
○ Subjects is willing to maintain their current exercise regimen and their current diet while on the study.

#### Exclusion criteria

○ Subjects who are participating in another Clinical Trial.
○ Subjects who faint during blood collection, or who have a phobia of needles, or a phobia of blood.
○ Subjects who are weightlifters.
○ Females who are pregnant or lactating (verbal confirmation only).
○ Subject who has taken any creatine supplements in the last 30 days.
○ Subjects with history of cancer (except localized skin cancer without metastases) within 5 years prior to the screening visit.
○ Subject has diabetes (insulin dependent or not) or any immune deficiency disease such as HIV/AIDS, etc.
○ Subject has any medical condition whether treating with mediation or not including hypertension (high blood pressure), hepatitis, any kidney disease or disorders, or musculoskeletal disorders. Note: if there are any significantly abnormal results for the BUN/creatinine screening labs the subject will be dropped.
○ Subject is taking a medication that would preclude participation in the study per the Principal Investigator.
○ Subjects with recent or current medical condition that may significantly impact the subject or the validity of the study results in the opinion of the Investigator.
○ Subjects who have a known or suspected allergy or sensitivity to creatine.
○ Chronic use of NSAIDs (e.g., Ibuprofen, Advil), or use of NSAIDs at one-week prior to each study visit.

#### Subject Withdrawal

The participation of a subject in this study may have been discontinued for any of the following reasons:

- the subject wishes to withdraw from study participation;
- if, in the opinion of the Investigator, it is in the best interest of the subject;
- suspected adverse effects from the test article;
- concurrent illness;
- violation of the prohibitions and restrictions (Section 5.3);
- development of an exclusion criteria. Subjects were free to withdraw at any time and need not give a reason, however, every reasonable attempt was made to ascertain such reasons. The data for those subjects who are withdrawn will be included in the final clinical report but may be excluded from final data analysis. Subjects were not followed up with after their withdrawal from the study, except in the case of a serious adverse event. Withdrawn subjects were not replaced.

#### Test Materials, Dosing and Randomization

Subjects were randomly assigned into one of 2 Groups, A or B according to the randomization (Protocol, Appendix 2) and were dosed accordingly.

Test Article Dosing

Group A (standard monohydrate group)

- 5g of creatine monohydrate after first meal of the day, once per day with the following instructions: After the first meal of the day subjects took 1 scoop of the test article powder mixed with 8 – 10 ounces water and stirred well.

Group B (monohydrate loading group)

- 20g of creatine monohydrate/day for the first 2 weeks, followed by a 5g dose after first meal of the day for the remainder of the study period with the following instructions: First 14 days, Loading Period: After the first meal of the day, subjects took 4 scoops of the test article powder mixed with at least 12-ounces water and stirred well. Next 14 days, Maintenance Period: After the first meal of the day subjects took 1 scoop of the test article powder mixed with 8 – 10-ounces water and stirred well.

#### Laboratory Analysis

All subjects had blood and urine analysis performed at baseline and the conclusion of the study. The laboratory tests included:

- CBC with differential
- Comprehensive Metabolic Panel
- Lipid Panel
- Creatinine, 24-Hour Urine
- Creatinine, Serum
- Creatine, Serum
- Amylase, Serum
- Lipase, Serum
- CK, Total + Isoenzymes, Serum

## Results

All subjects (n=24) completed the survey trial. Table 1 shows the demographic summary of test subjects by supplement dosage group. The demographics are similar for each group with the exception of no males in Group B. The urine and blood sample analysis results of the Baseline and Day 28 sample collections. showed that, under the study conditions, there were no safety concerns with 28 days of use of either of the two supplement doses (Creatine Monohydrate and Creatine Monohydrate with loading) tested in the study. There was one non-serious adverse event (AE) reported during the course of the study, menstrual cramps that the investigators considered moderate in severity and not related to the test article.

**Table 1.**
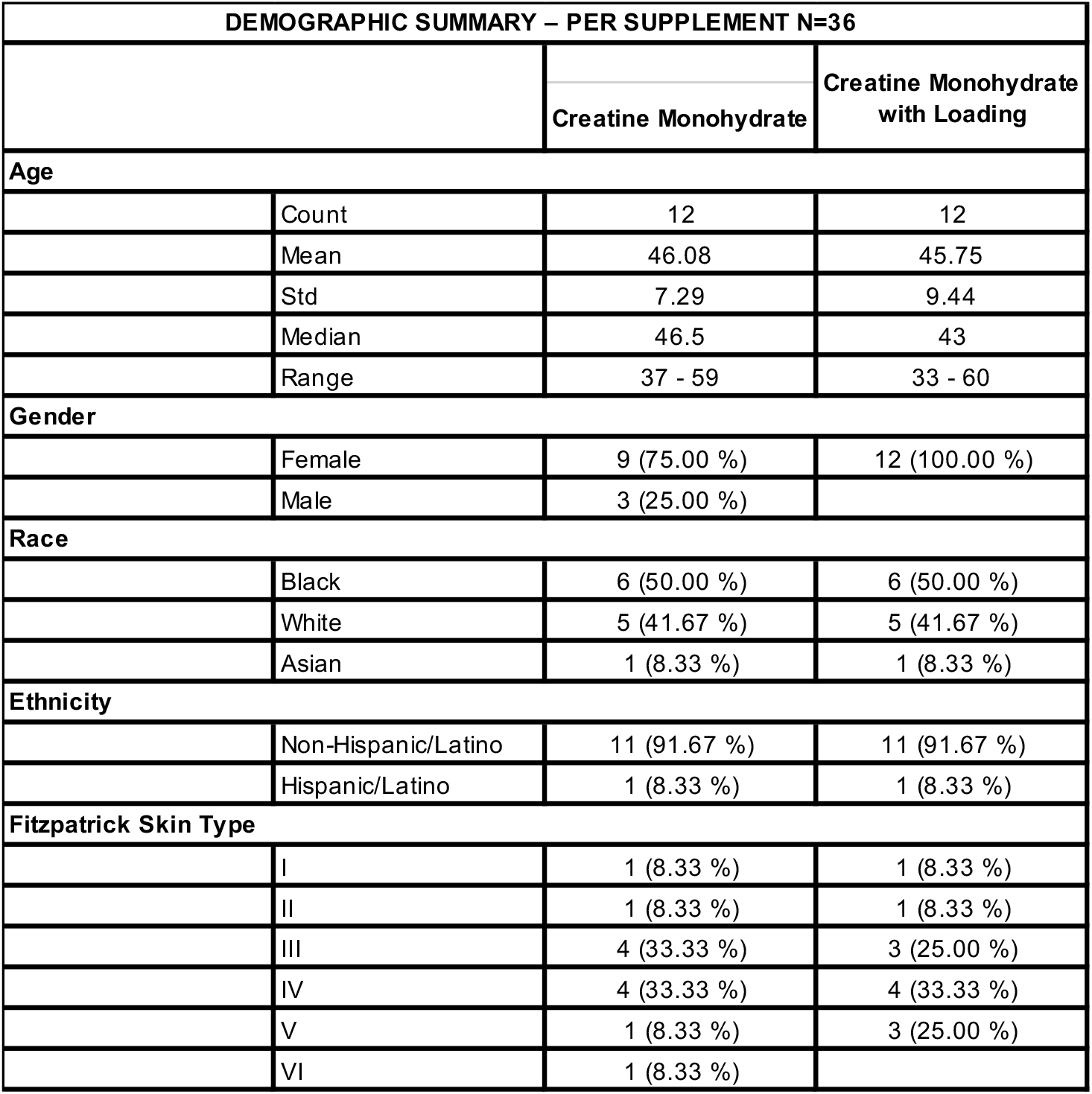
Demographic Summary of Test Subjects.

| DEMOGRAPHIC SUMMARY – PER SUPPLEMENT N=36 |  |  |  |
| --- | --- | --- | --- |
|  |  | Creatine Monohydrate | Creatine Monohydrate with Loading |
| **Age** |  |  |  |
|  | Count | 12 | 12 |
|  | Mean | 46.08 | 45.75 |
|  | Std | 7.29 | 9.44 |
|  | Median | 46.5 | 43 |
|  | Range | 37 - 59 | 33 - 60 |
| **Gender** |  |  |  |
|  | Female | 9 (75.00 %) | 12 (100.00 %) |
|  | Male | 3 (25.00 %) |  |
| **Race** |  |  |  |
|  | Black | 6 (50.00 %) | 6 (50.00 %) |
|  | White | 5 (41.67 %) | 5 (41.67 %) |
|  | Asian | 1 (8.33 %) | 1 (8.33 %) |
| **Ethnicity** |  |  |  |
|  | Non-Hispanic/Latino | 11 (91.67 %) | 11 (91.67 %) |
|  | Hispanic/Latino | 1 (8.33 %) | 1 (8.33 %) |
| **Fitzpatrick Skin Type** |  |  |  |
|  | I | 1 (8.33 %) | 1 (8.33 %) |
|  | II | 1 (8.33 %) | 1 (8.33 %) |
|  | III | 4 (33.33 %) | 3 (25.00 %) |
|  | IV | 4 (33.33 %) | 4 (33.33 %) |
|  | V | 1 (8.33 %) | 3 (25.00 %) |
|  | VI | 1 (8.33 %) |  |

Overall, 79.2% (19/24) of subjects in Groups A and B reported at least one GI symptom. Among women in both groups combined 81.0% (17/21) reported at least one GI symptom. In Group A the most common GI symptoms included water retention (50.0%), bloating, puffiness and weight gain (all at 41.67%). In Group B the most common GI symptoms were bloating (66.67%), stomach discomfort (58.33%0, water retention (50.0%), diarrhea and puffiness (both at 33.33%). Subjects in Group B rated puffiness, weight gain and stomach discomfort as more severe (16.67% vs 0.0%) compared to Group A. In addition to the trend of Group A reporting less severe GI and water retention symptoms, a significant difference was seen in Group A reporting no stomach discomfort as compared to Group B (Figure 1). Table 2 contains all of the subject responses regarding GI symptoms.

**Table 2.** Reported GI Symptoms by Severity.

|  | None |  | Mid |  |  |  |  |  |  |  | Severe |  |
| --- | --- | --- | --- | --- | --- | --- | --- | --- | --- | --- | --- | --- |
|  | 0 |  | 1 |  | 2 |  | 3 |  | 4 |  | 5 |  |
| Have you experienced: | Creat. MH | Creat. MH + L | Creat. MH | Creat. MH + L | Creat. MH | Creat. MH + L | Creat. MH | Creat. MH + L | Creat. MH | Creat. MH + L | Creat. MH | Creat. MH + L |
| 1. Bloating? | 58.33% | 33.33% | 25.00% | 33.33% | 8.33% | 0.00% | 0.00% | 16.67% | 8.33% | 8.33% | 0.00% | 8.33% |
| 2. Puffiness? | 58.33% | 66.67% | 16.67% | 8.33% | 16.67% | 0.00% | 8.33% | 8.33% | 0.00% | 8.33% | 0.00% | 8.33% |
| 3. Water retention? | 50.00% | 50.00% | 16.67% | 16.67% | 25.00% | 0.00% | 0.00% | 16.67% | 8.33% | 8.33% | 8.00% | 8.33% |
| 4. Weight gain? | 58.33% | 75.00% | 0.00% | 0.00% | 33.33% | 0.00% | 8.33% | 8.33% | 0.00% | 16.67% | 0.00% | 0.00% |
| 5. Stomach discomfort? | 83.33% | 41.67% | 16.67% | 33.33% | 0.00% | 0.00% | 0.00% | 8.33% | 0.00% | 16.67% | 0.00% | 0.00% |
| 6. G.I (gastro-intestinal) distress or diarrhea? | 91.67% | 66.67% | 0.00% | 16.67% | 0.00% | 8.33% | 0.00% | 0.00% | 0.00% | 8.33% | 0.00% | 0.00% |

**Fig. 1.**
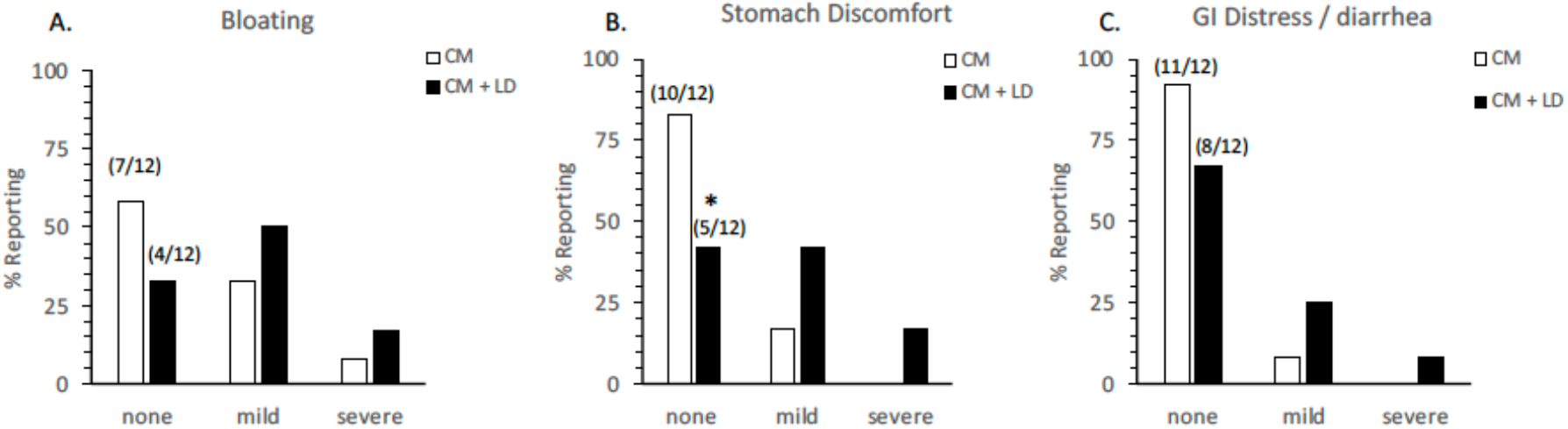
Statistical Analysis of GI Survey Results. Examination of survey results for Gl related side effects following 28-day CM supplementation. Study participants taking 5g daily dose of CM without a loading dose phase (CM) or following a 14-day, 20g loading dose (CM + LD) were evaluated for symptoms of Bloating (A), Stomach Discomfort (B) or Gl Distress and Diarrhea (C). Values represent the percent responding from each treatment group (n=12). * p < 0.05 compared to CM group based on Fisher’s Exact Test.

## Discussion

Creatine has been used by athletes since the early 1990s. Over time multiple studies have shown that creatine monohydrate (CM), the most common form of creatine, is both safe and effective. (10) As more individuals take creatine for performance and health benefits the anecdotal reports of GI symptoms and fluid retention persist. There is a theoretical basis for these symptoms associated with creatine monohydrate given its poor solubility of approximately 18mg/ml. (11)

The results of this study showed that male and female subjects experienced a variety of GI symptoms most notably bloating, water retention, stomach discomfort and puffiness. The incidence of symptoms between the two groups were not shown to be statistically significant.

However, there was a trend that Group B subjects experienced GI symptoms more frequently and reported those symptoms as more severe when compared to Group A. Group A subjects reported significantly less stomach discomfort as compared to Group B. To rule out potential gender related differences these data were re-analyzed with the male subjects removed. Even with the male subjects removed, there was a statistically significant increase in reported stomach discomfort in Group B.

This potential dose-related effect has been previously reported. In a 2008 study Ostojic and Ahmetovic looked at GI distress secondary to creatine supplementation for 28 days in male athletes. CM was dosed as either 2 x 5 gm (C5 cohort) or 1 x 10 gm (C10 cohort). They found that the GI complaints were infrequent, mild and similar to placebo. However, there was a significant difference in the incidence of diarrhea between the C5 and C10 groups suggesting a possible dose-related phenomenon. (6)

Dose-dependent differences have also been shown for the oral bioavailability of CM by Alraddadi et al. (11) In their animal trial the investigators examined the oral bioavailability of low and high doses of CM in rats as compared to IV administration of CM. The bioavailability of the low and high doses of CM was 53% and 16% respectively. The authors hypothesized the dose-dependent differences between the low and high CM doses may be due to a saturation of the intestinal transport driven processes or incomplete dissolution of the solid dosage forms. The low bioavailability of the higher CM dose runs contrary to what is frequently referenced.(12) The claims of complete 100% bioavailability for CM supplements are based on scant evidence from trials using tracer doses of CM much lower then typically used for dietary supplementation (less than 1 gm) as well as an absence of a IV CM administration comparator. (12,13)

Creatine monohydrate is currently being used as a treatment or adjuvant for neurological conditions using doses as high as 20 gm/day. (8) As higher doses of CM are employed tolerability becomes an issue and can impact compliance. Although mild, reversible GI symptoms are not considered a safety issue they may be sufficient to dissuade a patient from continuing use. Similarly, fluid retention and puffiness can negatively impact compliance due to performance and aesthetic effects. In a recent pilot study, researchers took tolerability into consideration where they found that 19 of 20 patients with Alzheimer’s disease taking CM 20gm/day (taken as two 10 gm doses) in the trial reached 80% compliance. (9) High dose creatine monohydrate has also been studied for other neurological conditions including sleep deprivation, traumatic brain injury and depression utilizing doses ranging from 10 gm/day to greater than 20 gm/day. (8) In the majority of these studies patients were dosed with CM for a period of time ranging from a single day to 8 weeks. For drug therapy, adherence is approximately 50% in patients with chronic disease. More frequent dosing, a common strategy for drugs with tolerability issues, results in an increased rate of noncompliance. (14)

The current study was designed to determine if two commonly used dosage regimens for creatine monohydrate produced gastrointestinal symptoms or other adverse events that would impact supplementation compliance. Limitations of the trial include the small number of subjects in each group which may limit generalizability. The use of self-reported outcomes potentially could introduce bias. The protocol did not control for GI symptoms related to menses in pre-menopausal female subjects. While creatine monohydrate has an extensive history as a dietary supplement for athletic performance it is heavily weighted toward male athletes and as such the potential frequency and intensity of GI related adverse events with CM use during menses has received less attention.

## Conclusion

Taking creatine monohydrate 5 gm/day with or without a loading dose resulted in a variety of gastrointestinal symptoms. Although not statistically significant a trend was observed that subjects in the loading dose group reported more symptoms that tended to be slightly more severe based on the rating scale. Future research considerations should include investigating the dose-related nature of GI symptoms of creatine monohydrate up to and including 20 gm/day, determining the impact GI symptoms have on compliance vs a control group and determining if novel creatine derivatives with superior solubility compare favorably to creatine monohydrate resulting in fewer and/or less severe GI symptoms.

## Data Availability

All data produced in the present study are available upon reasonable request to the authors

